# Brain age prediction improves the early detection of Alzheimer’s disease in East Asian elderly

**DOI:** 10.1101/2023.02.28.23286588

**Authors:** Uk-Su Choi, Jun Young Park, Jang Jae Lee, Kyu Yeong Choi, Sungho Won, Kun Ho Lee

**Author notes:** **Correspondence:** Kun Ho Lee, Gwangju Alzheimer’s & Related Dementia cohort research center and Department of Biomedical Science, Chosun University, Gwangju 61452, Republic of Korea. Contributed equally to this work (Uk-Su Choi, Jun Young Park).

## Abstract

**Introduction:** Brain age prediction is used to quantify the pathological and cognitive changes associated with brain aging. However, the predicted age derived from certain models can result in biased estimation and the concealment of inherent aged brain function.

**Methods:** We constructed a brain age prediction model for the East Asian elderly brain using the brain volume and cortical thickness features from cognitively normal (CN) brains. Furthermore, our model was used to estimate different diagnoses and to construct a classification model of mild cognitive impairment (MCI) conversion and Alzheimer’s disease (AD) conversion.

**Results:** Our model showed a strong association of the brain age difference (BAD) with three diagnosis groups. In addition, the classification models of MCI conversion and AD conversion showed acceptable and robust performances, respectively (area under the curve [AUC] = 0.66, AUC = 0.76).

**Discussion:** We believe that our model can be used to estimate the predicted status of an East Asian elderly brain. Moreover, the MCI conversion model has the potential to prevent severe cognitive impairment and can be used for the early detection of AD.

## Introduction

Biological age represents the individual age-related biological changes that are crucial for the evaluation of age-associated diseases that occur throughout life [1,2]. Age has been robustly studied because it is significantly related to mortality risk in humans [3]. However, biological changes cannot be linked to a single factor. In fact, biological changes are associated with multiple individual factors, such as genes, health, environment, and lifestyle. It can be estimated by the robustness of genetic changes, such as DNA methylation state [4,5] and individual lifestyle choices, such as eating style, sleep quality, and physical pattern [6]. Many age-related factors have been used to evaluate individual biological changes; therefore, various scientific approaches in diverse fields have been attempted. Among these approaches, one of the representative biological aging studies has focused on brain age. Brain age represents not only the current brain aging status but also the acceleration of brain aging [7,8] by using the difference between chronological age and predicted age [9]. These differences have been associated with a number of neurodevelopmental [10] and neurodegenerative diseases, such as Alzheimer’s disease (AD) and Parkinson’s disease [11].

Aging of the brain, including cognitive decline, is strongly associated with structural changes [12,13]. Magnetic resonance imaging (MRI) has been widely used to evaluate structural changes in the brain because it provides multiple contrasts between brain tissues and subcortical imaging in a noninvasive manner [14,15]. Atrophy of the cortical regions is one of the most robust age-related brain changes and leads to changes in the frontal and temporal regions of gray matter [16] and white matter [17] in elderly brains. In contrast to the normal brain aging process, the brain atrophy rate can also be slower according to cognitive abilities, such as in SuperAgers [12]. Several brain regions, such as the amygdala and hippocampus [18,19] do not fully support normal aging processing in the brain; rather, they are more closely related to neurodegenerative diseases, such as mild cognitive impairment (MCI) or Alzheimer’s disease (AD).

Most previous studies have focused on brain atrophy with respect to the cortical volume and thickness. These measurements represent brain atrophy and are comparable to each other, but show slightly different structural aspects [20,21]. Volume estimation can be conducted in both the cortical and subcortical regions, whereas thickness estimation is not possible in subcortical regions, such as the amygdala and hippocampus [22]. In addition, volume measures are commonly used as normalized values corrected by intracranial volume (ICV) due to the different brain sizes of subjects; however, cortical thickness should be used as absolute values (mm) [23]. Different genetic contributions to cortical volume and thickness can also support discrete structural aspects [24]. In age-related volume studies, ventricular volume changes were strongly associated with age [19]. Decreased posterior cortical volume resulting from visual dysfunction may not be directly associated with AD [25]. In age-related thickness studies, age-related decreases in the ratios of cortical thickness in the occipital and temporal lobes [26] and large subject variability in frontal and temporal areas were identified [27]. Thus, both cortical volume and thickness measurements may contribute to brain age estimations.

Here, we calculated both the brain structural volume and cortical thickness from 1,690 elderly East Asian brains and constructed a brain age prediction model based on the calculated measurements. We measured the brain age difference (BAD) from the prediction model and applied BADs to investigate the association with different diagnosed groups and the classification of conversion groups (MCI conversion and AD conversion) with our longitudinal dataset.

## Methods

### Subjects for brain age prediction model training

This study was approved by the Institutional Review Board of Chosun University Hospital, and all participants provided written informed consent. A total of 2,381 participants were recruited from Gwangju, Republic of Korea, and participated in this study. The study group consisted of 1,690 cognitively normal (CN), 476 MCI, and 215 AD subjects. All subjects were screened using several neuropsychological tests, including the Korean version of the Mini-Mental State Examination (K-MMSE) [28], the Seoul Neuropsychological Screening Battery (SNSB) [29], and the Clinical Dementia Rating (CDR) [30]. It was confirmed that the CN subjects had no history of neurological or psychiatric disorders, or impairments in daily activities.

### Subjects for classification model of conversion groups

In our dataset, different conversion groups were recruited from Gwangju, Republic of Korea, to participate in this study. A total of 364 and 239 participants who were diagnosed as CN and MCI at baseline, respectively, had at least one follow-up (mean follow-up duration (years):3.0/2.5 (CN/MCI)). During the follow-up period, 84 CN and 47 MCI subjects were converted to MCI and AD, respectively, based on a physician’s final decision. We divided the subjects into conversion (CNc, converted CN; MCIc, converted MCI) and non-conversion groups (CNs, stable CN; MCIs, stable MCI).

### MRI acquisitions

All structural MRI images of the Asian cohort data were acquired using 3.0 T (Skyra, Siemens). The T1-MPRAGE sequence was acquired with the following parameters: TR = 2300 ms, TE = 2.143 ms, TI = 900 ms, FOV = 256 × 256, matrix size = 320 × 320, thickness = 0.8 mm. The T2-SPACE sequence was acquired with the following parameters: TR = 2300 ms, TE = 2.143 ms, TI = 900 ms, FOV = 256 × 256, matrix size = 320 × 320, thickness = 0.8 mm.

### Calculation of structural brain volumes and cortical thickness

All T1 images were analyzed using Neuro I (V.1.3; http://neurozen.ai/), which is a commercial software for neuroimaging analysis. The software package calculated structural brain volumes and cortical thickness as follows: (1) T1 structural images were corrected for bias field inhomogeneity using N4 [31]. (2) The corrected images were skull-stripped using a deep learning model based on a 3D convolution neural network. (3) The extracted brain images were parcellated into 107 regions of interest (ROIs) based on the Desikan-Killiany-Tourville (DKT) atlas using a 3D CNN deep learning model. The cortical and subcortical volumes were measured by normalizing the voxel numbers of the parcellated ROIs by the intracranial volume (ICV). The cortical thickness of parcellated ROIs was calculated using a project-based thickness (PBT) algorithm [32].

### Brain age prediction model of CN group using linear regression

We constructed a brain age prediction model with linear Elastic Net Regression using 1,690 CN subjects. With regards to the predictor, the calculated cortical thickness and subcortical volume measures from all parcellated ROIs and sex were considered. We optimized several hyperparameters of the prediction model by maximizing the prediction accuracy using the grid search method.

The performance of the model was evaluated in two ways. First, five-fold cross-validation (CV) was adopted by splitting the folds in such a way that each fold contained a similar distribution for the age bin. CV accuracy was calculated by averaging the five-fold validation accuracy. The metrics used were the mean absolute error (MAE) and Pearson correlation (*r*) between chronological age and predicted age. Second, 476 MCI subjects and 215 AD subjects were adjusted to the final brain age model, and their predicted ages were compared with those of the CN subjects. All statistical analyses were conducted using the Scikit-learn package (version 1.2.0) in Python (version 3.7).

### Brain Age Difference (BAD) calculation

To evaluate the acceleration of brain aging, we calculated the differences between chronological age and the predicted age derived from our prediction model, which was referred to as brain age differences (BADs). The chronological age, predicted age, and BAD for the *i*th subject was denoted as *y*_*i*_, *ŷ*_*i*_ and *BAD*_*i*_, respectively. Thus, the BAD for the *i*th subject can be represented as follows:

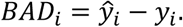

### Association of BAD with different diagnosis groups

To determine whether BADs differed by group (CN vs. MCI vs. AD), we conducted Analysis of Variance (ANOVA) and Analysis of Covariance (ANCOVA) adjusting for sex and age as covariates in order to confirm the differences in BADs by conversion groups (CNs vs. CNc and MCIs vs. MCIc).

### The classification model of conversion groups (MCI and AD)

To prepare accurate conversion group data for the training of the classification model, we selected conversion-type data at the restricted time point that was robustly represented. Among the follow-up data of the conversion groups (CNc and MCIc), the data at the time point right before conversion to MCI or AD were considered, and the data of the non-conversion groups (CNs and MCIs) at the time point right before the last follow-up time were selected for a fair comparison with the conversion groups. Using these data, we developed three classification models. First, the baseline model was built using chronological age, sex, and years of education. Second, the MMSE scores were added to the baseline model. Third, we considered demographic characteristics (age and sex) and BAD as covariates. All combinations of covariates were adjusted to predict the conversion of MCI and AD, and all models were built using logistic regression.

To evaluate the performance of the models, a five-fold CV was adopted, and the metrics used were the area under the ROC curve (AUC), sensitivity, and specificity. To compare the AUCs for different covariates, we conducted the Mann–Whitney U-test [33,34]. We conducted all analyses with Scikit-learn package (V.1.2.0) and Scipy (V.1.9.3) in Python (V.3.7).

## Results

### Descriptive statistics of study subjects

Table 1 shows the demographic characteristics of the study sample. First, 2,381 subjects were used to develop and evaluate the brain age models. The subjects were allocated to one of three groups (1,680 CN subjects, 476 MCI subjects, and 215 AD subjects) (Table 1A). The average ages were 71.3 (± 6.9) years, 73.2 (±6.8) years and 75.4 (±7.2) years for CN, MCI and AD, respectively. The education levels were similar between CN (10.2±4.5) and MCI (10.4±4.6), while AD subjects had significantly lower education levels (p < 0.01). The MMSE scores differed significantly among the groups and the lowest means were observed for subjects with AD (CN: 27.1±2.3; MCI: 25.3±2.9; AD: 19.4±5.5). The demographic information for the conversion groups is shown in Table 1B. Among the 364 CN subjects at baseline, 84 subjects were converted to MCI subjects (CNc), while 280 subjects remained (CNs). The mean follow-up period was 3.1±1.9 years and 2.5±1.9 years for CNs and CNc, respectively. Approximately 239 MCI subjects at baseline consisted of 47 subjects who were converted to AD (MCIc) and 192 subjects who were still diagnosed as CN (MCIs). The mean follow-up period was 2.5±1.5 years and 2.4±2.1 years for MCIs and MCIc, respectively. The mean ages of the conversion groups were higher than those of the non-conversion groups in both CN and MCI (CNs:71.0±5.1 years; CNc:73.3±5.7 years; MCIs:72.1±5.3 years; 75.2±6.4 years). There was no significant difference in the education levels and MMSE scores between CNs and CNc; however, the education levels and MMSE scores for MCIs and MCIs differed significantly (p < 0.01).

**Table 1:** Demographic information.

| Group | Total | CN | MCI | AD |
| --- | --- | --- | --- | --- |
| N | 2,381 | 1,690 | 476 | 215 |
| Age, mean, years (SD) | 72.0 (6.9) | 71.3 (6.7) | 73.2 (6.8) | 75.4 (7.2) |
| Male sex, N (%) | 988 (41.5) | 655 (38.8) | 237 (33.4) | 96 (44.7) |
| Education, mean, years (SD) | 10.0 (4.6) | 10.2 (4.5) | 10.4 (4.6) | 8.1 (5.0) |
| MMSE score mean, (SD) | 26.0 (3.6) | 27.1 (2.3) | 25.3 (2.9) | 19.4 (5.5) |

MMSE: Mini-Mental State Examination
SD: Standard deviation

| Group | Non Conversion |  | Conversion |  |
| --- | --- | --- | --- | --- |
|  | CNs | MCIs | CNc | MCIc |
| N | 280 | 192 | 84 | 47 |
| Baseline age, mean, years (SD) | 71.0 (5.1) | 72.1 (5.3) | 73.3 (5.7) | 75.2 (6.4) |
| Male sex, N (%) | 123 (43.9) | 98 (51.0) | 40 (47.6) | 26 (55.3) |
| Education, mean, years (SD) | 10.9 (4.3) | 10.6 (4.7) | 11.0 (4.5) | 8.5 (4.5) |
| MMSE score mean, (SD) | 27.9 (1.8) | 25.9 (2.8) | 27.3 (2.3) | 23.2 (3.2) |
| Follow-up period, mean, years (SD) | 3.1 (1.9) | 2.5 (1.5) | 2.5 (1.9) | 2.4 (2.1) |

CNs: CN stable; MCIs: MCI stable; CNc: converted to MCI; MCIc: converted to AD

### Brain age prediction model of CN group using linear regression

We adopted the linear ElasticNet regression algorithm to acquire the best baseline performance and optimized hyperparameters for the final model. Our model exhibited outperformance (r = 0.803, MAE = 3.25 years) when compared with previous models [35,36] (Fig. 1A). The BADs for our age prediction model exhibited a negative correlation (r = -0.06), as reported in previous studies [37,38].

**Figure 1:**
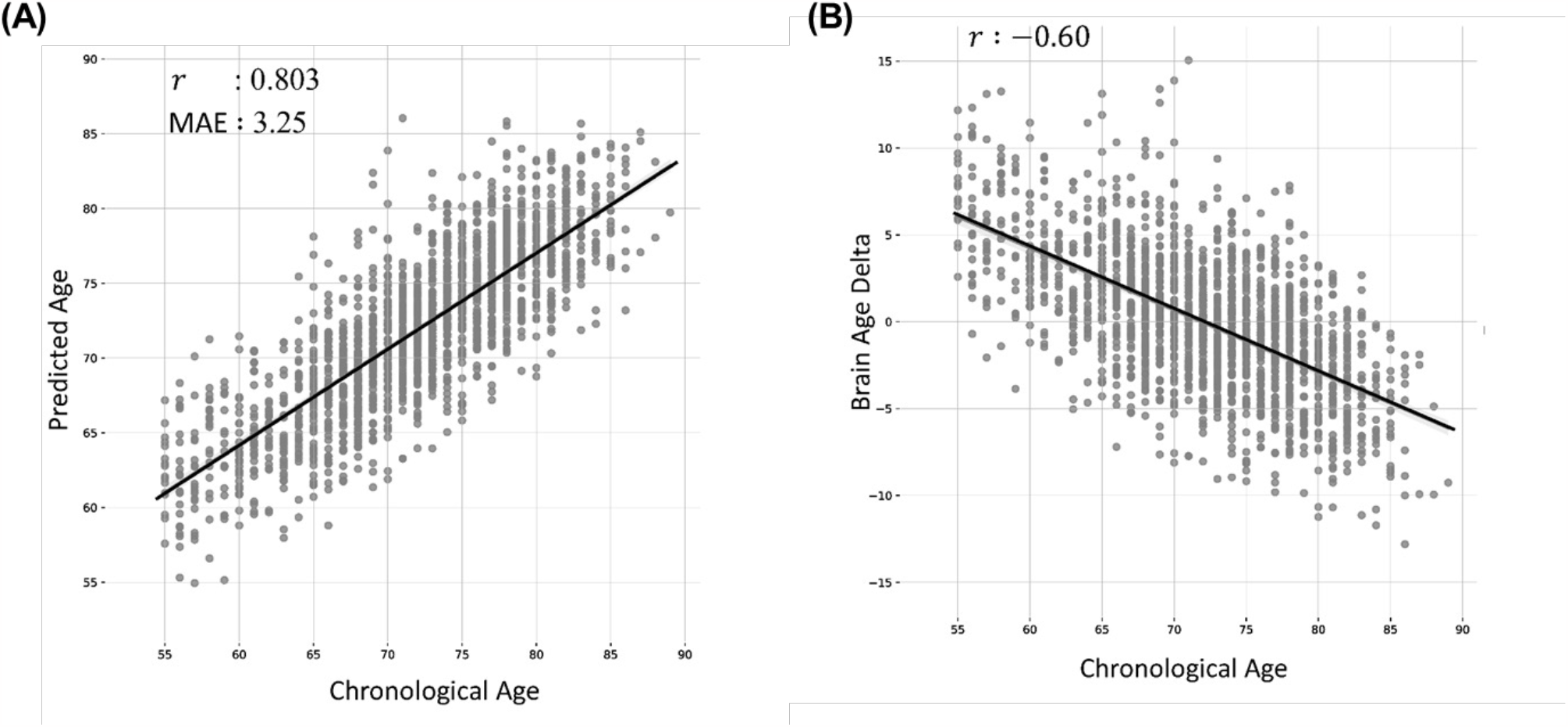
Brain age prediction model. (A) the prediction performance of the proposed model. (B) Brain age delta derived from the proposed model.

### Association of BAD with different diagnosis group

We evaluated three different groups (CN, MCI, and AD) using the BAD measurements. All groups presented significantly different BADs (p < 0.001, ANCOVA). The AD group represented the highest intercept (43 years) and the lowest slope (r = 0.43) among all three groups (r = 0.64 and intercept = 25.7 years for CN; r = 0.66 and intercept = 26.6 years for MCI) (Fig. 2A). Interestingly, the trend line of the AD group met that of the MCI group at 89.0 years of age. The mean BADs values of the CN, MCI, and AD groups were 0.01 years, 1.5 years and 3.8 years, respectively. The AD group had larger BAD values than the MCI and CN groups (p < 0.001 for AD vs. CN; p < 0.001 for AD vs. MCI). In addition, the MCI group had a larger BAD than the CN group (p < 0.001) (Fig. 2B).

**Figure 2:**
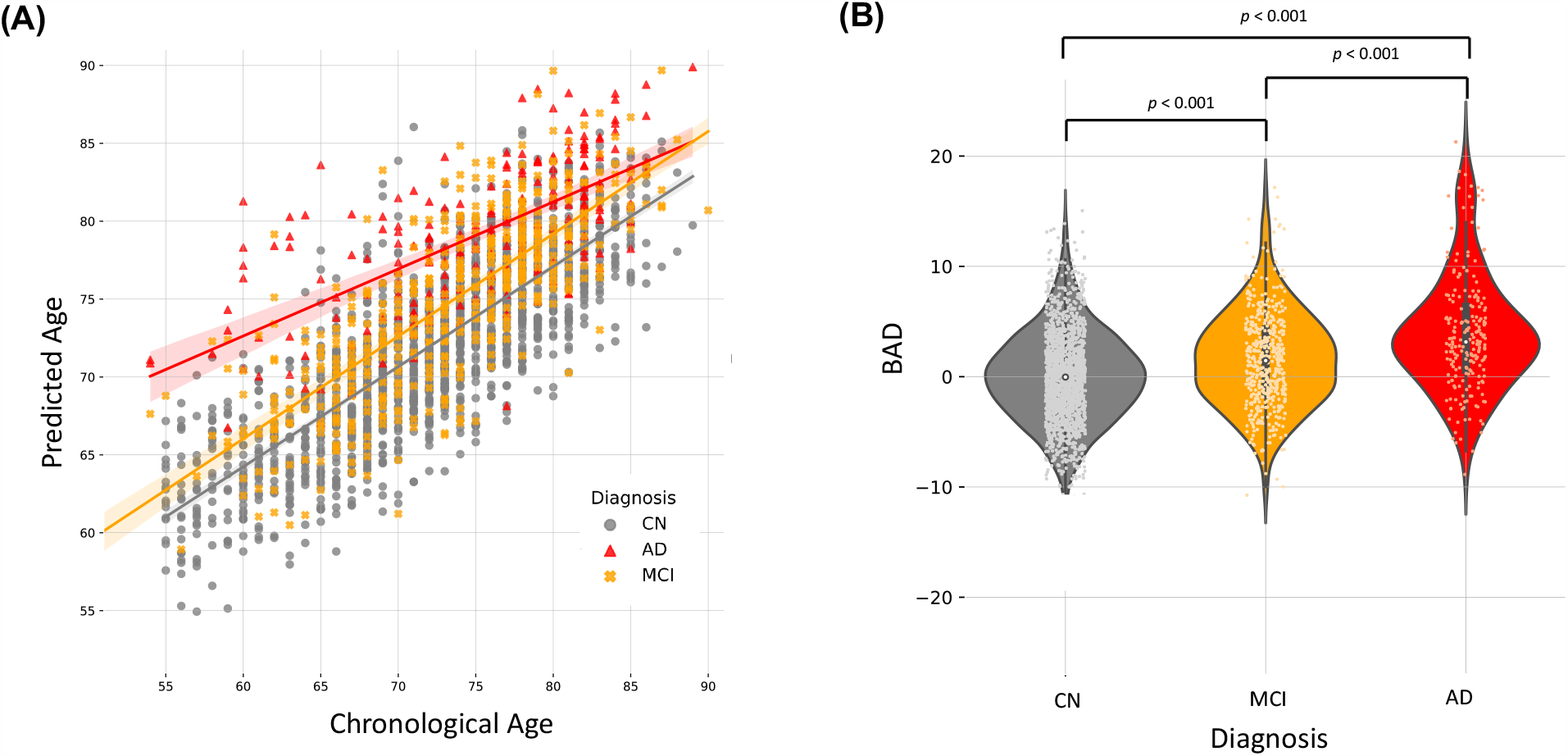
Brain age prediction of different diagnosis groups; Cognitively normal (CN), mild cognitive impairment (MCI), and Alzheimer’s Disease (AD). (A) the prediction of CN, MCI, and AD. (B) Brain age differences (BADs) of CN, MCI, and AD.

### The classification model of conversion groups

The non-conversion groups (CNs and MCIs) had significantly different BADs than the conversion groups (CNc and MCIc) when sex and age were adjusted as covariates (p = 0.005 for CNs versus CNc and p < 0.001 for MCIs versus MCIc) (Fig. 3). Based on these findings, we constructed classification models to predict conversion to MCI and AD. The proposed classification models of MCI conversion demonstrated an acceptable performance (AUC = 0.66, sensitivity = 0.62, specificity = 0.59), and AD conversion demonstrated a robust performance (AUC = 0.76, sensitivity = 0.66, specificity = 0.71). Furthermore, the proposed model of MCI conversion and AD conversion exhibited a significantly higher AUC than the baseline models due to education level (p < 0.01 for MCI conversion; p < 0.01 for AD conversion) (Fig. 4).

**Figure 3:**
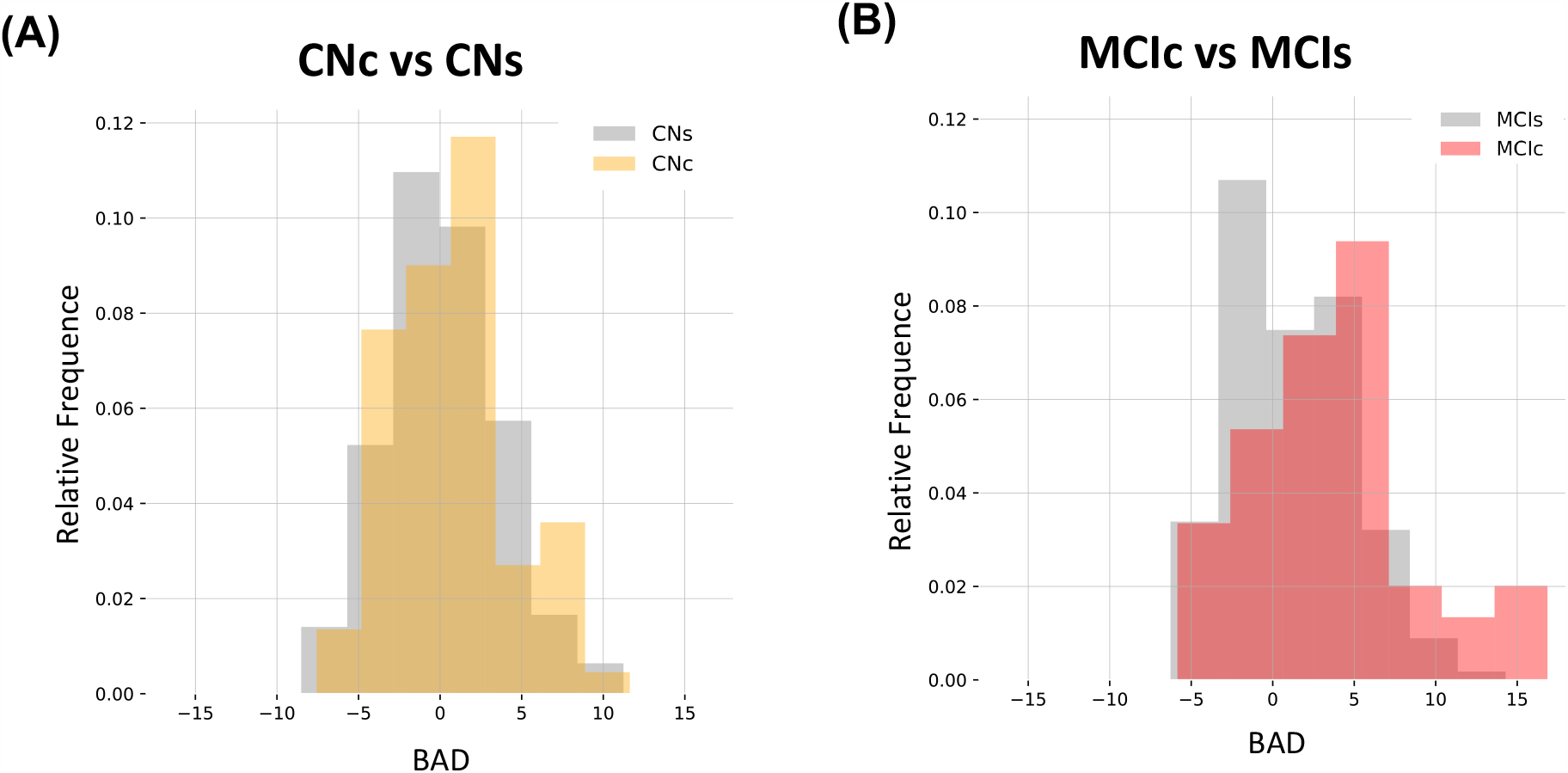
Brain age prediction and brain age delta (BAD) distribution of different conversion groups. (A) The distribution of CN converted (CNc) and CN stable (CNs) groups. (B) The distribution of MCI converted (MCIc) and MCI stable (MCIs) groups.

**Figure 4:**
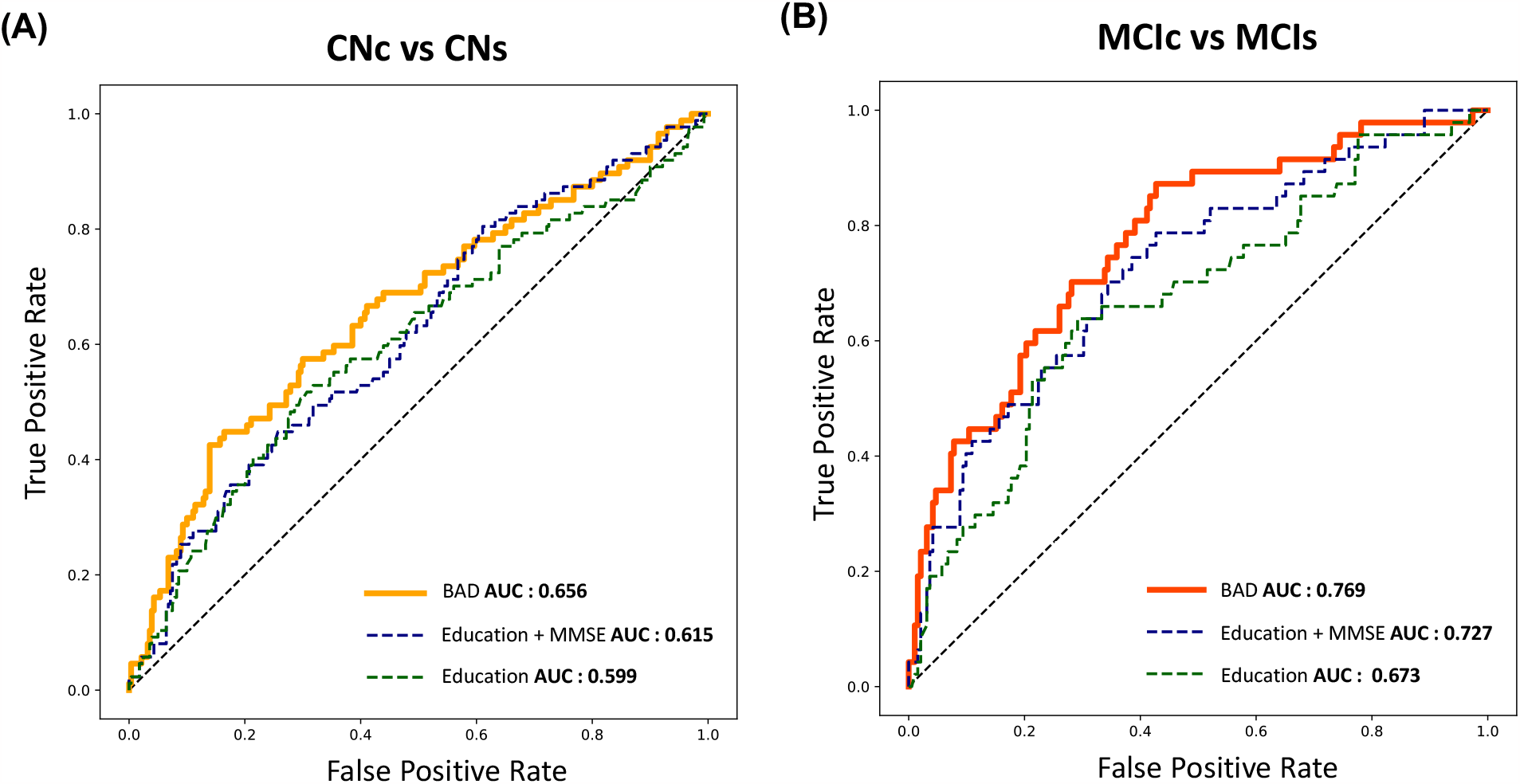
MCI and AD conversion classification models. (A) Area under the curve (AUC) plot of the MCI conversion classification model. (B) AUC plot of the AD conversion classification model.

## Discussion

This study demonstrated a robust brain age prediction (r = 0.803, MAE = 3.25 years) of elderly East Asian brains. The BADs derived from our model had a strong association with different diagnosis groups (p < 0.001, ANCOVA). Our longitudinal data showed significant differences in BADs between the conversion groups (CNc and MCIc) and non-conversion groups (CNs and MCIs). Furthermore, the differences in BADs resulted in the robust performances of the classification models between conversion and non-conversion groups (AUC = 0.66 for MCI conversion, AUC = 0.77 for AD conversion, respectively).

Previous brain age prediction models have been trained using a lifespan dataset with a wide range of ages and demonstrated an accurate prediction with minimal errors [39,40]. These models commonly adopted MRI-based biomarkers, such as brain volume or cortical thickness of parcellated ROIs [36] derived from T1w images. Nonetheless, a recent study has reported a more accurate brain age prediction model using a multimodal strategy [41].

However, these types of models have been found to be unsuitable for the evaluation of narrow age ranges, such as the younger or elderly groups. Several studies have reported the bias estimation of different age range groups, underestimation of younger ages, and overestimation of older age due to the systematic bias of inherent regression methods [11,42]. In fact, our model also demonstrated an inherent bias in brain age prediction in elderly brains (Fig. 1B). Until recently, there have been very few datasets available for a specific range of ages due to difficulties related to data acquisition, with the exception of a specific consortium, such as the Alzheimer’s Disease Neuroimaging Initiative (https://adni.loni.usc.edu/).

To overcome the limitations of previous approaches, our brain age prediction model was constructed using a large cohort dataset of elderly East Asian brains. The proposed prediction model had a smaller error (MAE = 3.25) and a larger correlation coefficient (r = 0.803) than the other models [35,36]. Furthermore, the trend lines and BADs derived from our model had a strong association with the different diagnosis groups (p < 0.001) (Fig. 2B). Interestingly, the crossover of AD and MCI trend lines at older ages (Fig. 2A) strongly supports the notion that chronological age is a key factor in AD [43]. In addition, the BADs also indicated an association with differences between the conversion and non-conversion groups (p < 0.01, MCI conversion; p < 0.001, AD conversion) (Fig. 3). Previous studies have shown that the trajectory of diagnosis changes from MCI; however, most of these studies were based on brain atrophy, cognitive measurements, and positron emission tomography features [44,45]. The BAD derived from our brain age prediction model has great potential to elucidate intrinsic brain acceleration associated with neurodegenerative diseases [39]. In addition, our classification of MCI conversion (CNs versus CNc) is important for evaluating BADs as a biomarker for early cognitive impairment, since a large cohort of longitudinal CN datasets that can be divided into conversion and non-conversion types are difficult to acquire. However, our longitudinal datasets comprised large cohort data of elderly people, including both the conversion and non-conversion types of CN and MCI groups with fixed MRI acquisition protocols. Based on the classification results, the proposed model demonstrated acceptable classification performance for MCI conversion and robust performance for AD conversion (Table 2 and Fig. 4). The MCI conversion classification model demonstrated relatively lower accuracy due to the small sample size and differences in neurodegeneration between CNs and CNc [46].

**Table 2:** Prediction results for the classification of conversion groups. The baseline model was considered using chronological age, sex, and education. Other models were statistically compared with the baseline model to evaluate AUC differences using the Mann–Whitney U-test.

| Input modality | CNs vs CNc |  | MCIs vs MCIC |  |
| --- | --- | --- | --- | --- |
|  | AUC | *p-value* | AUC | *p-value* |
| Education | 0.599 [0.596 – 0.602] | - | 0.673 [0.670-0.675] | - |
| Education + MMSE | 0.615 [0.612 - 0.618] | <0.05 | 0.727 [0.725-0.730] | <0.01 |
| Brain Age | 0.656 [0.655 - 0.657] | <0.01 | 0.763 [0.760-0.772] | <0.01 |

Despite these findings, this study had several limitations. First, the small sample size of CNs weakened our novel classification model. To use our model for clinical applications, it needs a higher accuracy and sensitivity of at least 0.8 AUC [47]. Second, the suggested brain age prediction model was based on a single feature, namely the brain volume of the parcellated ROIs. Several studies have shown a higher AUC and lower MAE through multimodal features such as diffusion-weighted images, T1-weighted images, or genetic features [44,45]. Our model can be enhanced by using these multimodal features. Lastly, we did not evaluate our model using an external dataset with different ethnicities and MRI acquisition parameters. The shape and volume of the Caucasian brain are substantially different from those of the Asian brain, which can result in prediction errors [48] and different MR acquisition parameters can change the contrast of images, resulting in a slightly altered segmentation [49]. Thus, our model cannot be generalized, but the MRI acquisition parameters used in this study have been commonly used in the clinical field.

Despite these limitations, our study has great potential in shedding light on Alzheimer’s disease research. We constructed an elderly specific model of brain age prediction using a large East Asian cohort dataset, in contrast to other studies that have mostly adopted the ADNI dataset composed of Caucasians. The Asian brain has different aspects of morphology, and ethnicity-specific approaches can provide more accurate predictions [50]. To the best of our knowledge, we have proposed a novel MCI conversion classification model for elderly East Asian brains. In addition, MCI convergence from CN is hardly detected because of its small detectable morphological changes, cognitive changes, and reversion from MCI to CN due to cognitive rehabilitation [51,52]. These inherent limits of the datasets resulted in a lower performance of MCI conversion than that of AD conversion (Fig. 4). Although there have been a few studies [53,54] that indicated convergence from CN to MCI, our model can be applied to the early detection of AD prediction, especially in East Asian people.

In conclusion, we found that the BAD of our proposed brain age prediction model is a good candidate biomarker for cognitive impairment and neurodegenerative disease diagnosis. The novel classification model based on BAD demonstrated great potential for early AD detection and prediction, and can be useful for the clinical treatment of AD in the future.

## Data Availability

All data produced in the present study are available upon reasonable request to the authors

## Author contributions

U-SC and JP: conceptualization, data curation, methodology, investigation, visualization, and writing of the original draft. KL: funding acquisition, supervision, and project administration. KC and JL: resources. U-SC and JP: Writing, review, and editing. All authors have contributed to the manuscript and approved the submitted version.

## Availability of data and materials

The dataset for the current study is not publicly available but is available from the corresponding author upon reasonable request.

## Ethics approval and consent to participate

This study was approved by the Institutional Review Boards of Chosun University Hospital (CHOSUN 2013– 12–018-068) and Chonnam National University (CNUH-2019-279). Written informed consent was obtained from each participant or their legal guardian.

## Consent for publication

Not applicable.

